# Automatic Detection of Target Engagement in Transcutaneous Cervical Vagal Nerve Stimulation for Traumatic Stress Triggers

**DOI:** 10.1101/2020.01.27.20018689

**Authors:** Nil Z. Gurel, Matthew T. Wittbrodt, Hewon Jung, Stacy L. Ladd, Amit J. Shah, Viola Vaccarino, J. Douglas Bremner, Omer T. Inan

**Affiliations:** School of Electrical and Computer Engineering, Georgia Institute of Technology; Department of Psychiatry and Behavioral Sciences, Emory University School of Medicine; Department of Epidemiology, Rollins School of Public Health; Department of Cardiology, Emory University School of Medicine; Departments of Psychiatry and Behavioral Sciences and Radiology, Emory University School of Medicine; School of Electrical and Computer Engineering, Coulter Department of Biomedical Engineering, Georgia Institute of Technology

**Keywords:** traumatic stress, physiological biomarkers, vagal nerve stimulation, wearable bioelectronic medicine, wearable neuromodulation, wearable sensing

## Abstract

Transcutaneous cervical vagal nerve stimulation (tcVNS) devices are attractive alternatives to surgical implants, and can be applied for a number of conditions in ambulatory settings, including stress-related neuropsychiatric disorders. Transferring tcVNS technologies to at-home settings brings challenges associated with the assessment of therapy response. The ability to accurately detect whether tcVNS has been effectively delivered in a remote setting such as the home has never been investigated. We designed and conducted a study in which 12 human subjects received active tcVNS and 14 received sham stimulation in tandem with traumatic stress, and measured continuous cardiopulmonary signals including the electrocardiogram (ECG), photoplethysmogram (PPG), seismocardiogram (SCG), and respiratory effort (RSP). We extracted physiological parameters related to autonomic nervous system activity, and created a feature set from these parameters to: 1) detect active (vs. sham) tcVNS stimulation presence with machine learning methods, and 2) determine which sensing modalities and features provide the most salient markers of tcVNS-based changes in physiological signals. Heart rate (ECG), vasomotor activity (PPG), and pulse arrival time (ECG+PPG) provided sufficient information to determine target engagement (compared to sham) in addition to other combinations of sensors. resulting in 96% accuracy, precision, and recall with a receiver operator characteristics area of 0.96. Two commonly utilized sensing modalities (ECG and PPG) that are suitable for home use can provide useful information on therapy response for tcVNS. The methods presented herein could be deployed in wearable devices to quantify adherence for at-home use of tcVNS technologies.

## I. Introduction

THE vagus nerve is a complex cranial nerve which, via connections to the brain, neck, heart, lungs, and abdomen, mediates autonomic tone within the body. Efferent vagal projections interface with peripheral organs to mediate autonomic, endocrine, and behavioral responses [1]. Electrical stimulation of the vagus nerve has promising therapeutic effects on inflammatory, psychiatric, and cardiovascular disorders, either through implantable or noninvasive devices [2, 3]. The latter method includes transcutaneous stimulation of the auricular branch (in the ear) or the cervical branch (at the neck). Auricular and cervical VNS (taVNS and tcVNS) appear to activate cortical brain areas receiving vagal projections [4] along with limbic structures [5] which have been hypothesized to affect baroreception and prioprioception. Noninvasive vagal stimulation technologies have similar therapeutic benefits compared to the implanted counterparts, while avoiding the need for surgery. These noninvasive technologies have great potential for daily at-home use for rehabilitation, mood, and performance improvement [6, 7]. Moreover, existing research demonstrates their safety and tolerability for use in human subjects [8, 9].

Although taVNS and tcVNS have similar effects in imaging studies, they should be considered separate as the stimulation targets different locations of the vagus nerve -auricular branch through the ear and cervical branch through the neck, respectively. Hour-long sessions of taVNS have been shown to improve vagal tone [10, 11]. Favorable outcomes have been reported for migraine [12], epilepsy [13], major depression [14, 15], posttraumatic stress disorder [16]. Pairing taVNS with acute psychological stress was also studied: taVNS was shown to improve subjective fear and worry responses [17-19]. The newer and less common approach, tcVNS, was shown to decrease serum cytokines, and improve cardiac vagal tone [20, 21]. Clinically-relevant effects of minute-long sessions of tcVNS have been reported for conditions such as trigeminal allodyna [22], and cluster headache [23]. tcVNS is particularly suitable for acute therapies due to short session time (two minutes) when stress-related neuropathways are activated.

The investigation of tcVNS applied in tandem with acute traumatic stress is clinically relevant. Patients with trauma-related psychiatric disorders may experience traumatic flashbacks multiple times a day, triggered by, for example, sensory data (odor, sound, room size), cues related to the traumatic event, or even temporal data (incident repeated at the same time of the day) [24-28]. These specific triggers may be relevant to personal traumatic memories from sexual assault to combat exposure [29]. Recently, we demonstrated clinically-relevant physiological effects of tcVNS as measured by downstream physiological signals [30], as a first step to evaluate the merit of tcVNS usage in context of traumatic stress. Engineering efforts to transfer tcVNS to at-home use bring challenges regarding the stimulation application. A fundamental challenge is the determination of whether stimulation delivered properly [31]: The electrode placement and stimulation intensity affect the current flow and distribution, hence the stimulation efficacy, as tcVNS electrodes are directly in contact with the skin rather than placed on the nerve itself (as with an implantable counterpart). *Thus, personalized neuromodulation to provide feedback for the physician or for the patient on whether stimulation has been properly delivered to the vagus nerve is necessary for longitudinal treatment paradigms*. As summarized in Fig. 1, interfacing tcVNS with noninvasive sensing modalities and extracting information to decode whether the stimulation engaged the nerve target could dramatically improve stimulation efficacy and provide closed-loop delivery in response to a detected event. Previous work from our group has demonstrated that certain physiological parameters appear to be reflective of tcVNS delivery compared to sham [30, 32, 33], but the ability to accurately detect whether tcVNS has been delivered has never been investigated. Continuous cardiovascular and peripheral physiological sensing offer a convenient tool for this investigation, due to the intimate relationship of the vagus with the heart and the peripheral physiology, and the ubiquity of noninvasive wearable technologies. Previously, changes in individual features such as heart rate (HR) have been observed, the differences between groups in these individual features were not sufficient to allow for classifying whether or not tcVNS was properly delivered. Another point to consider is the optimization of the acquired modalities: while wearable sensing offers a convenient tool to quantify tcVNS, learning from the optimized modalities (i.e. “learning from less data”) would be favorable for hardware adaptations that possibly may not employ all measurement modalities discussed.

**Fig. 1.**
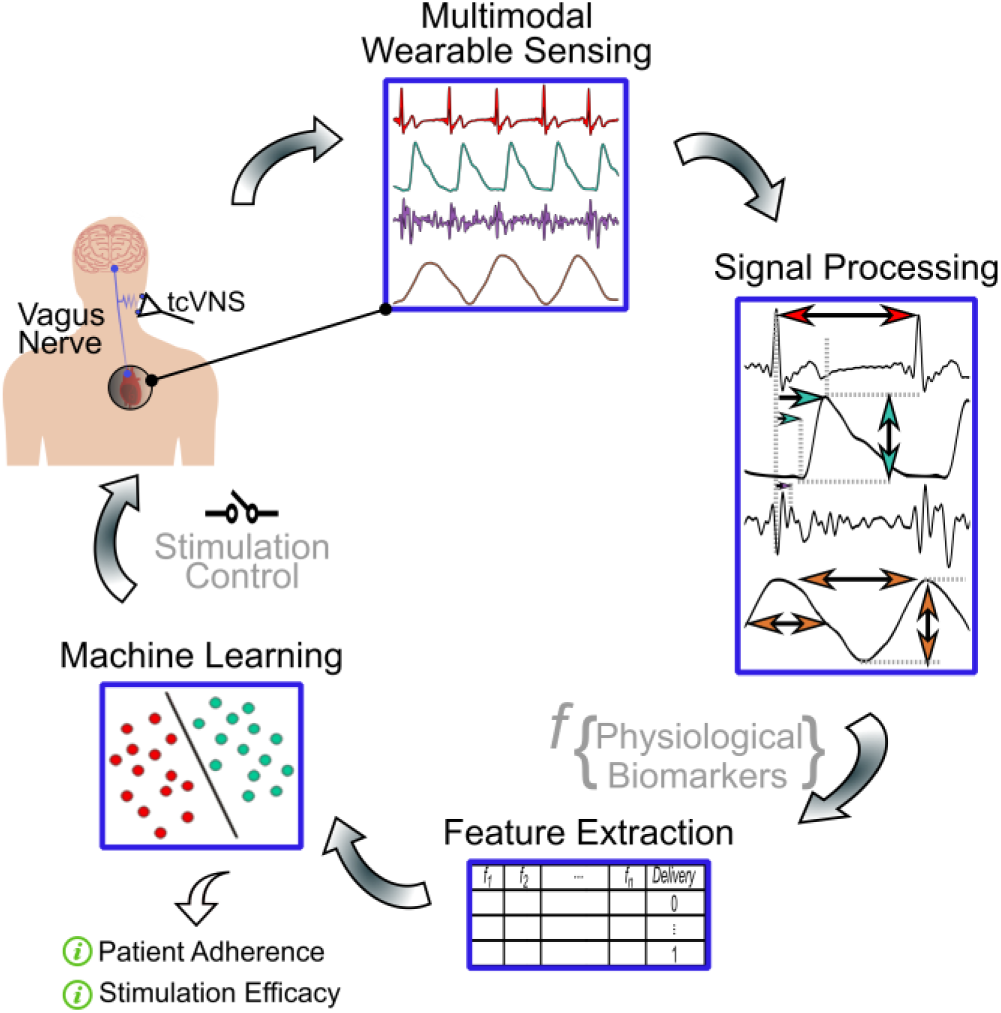
Wearable neuromodulation technologies can interface with noninvasive sensing. Ongoing activity acquired from one or more sensors can be utilized to assess cardiovascular and peripheral physiological response via signal processing. Events or changes in these data streams can then be decoded using machine learning via feature extraction, in order to dynamically trigger closed-loop delivery when needed, and to quantify patient adherence and stimulation efficacy for at-home therapy.

In this work, we conducted a study with 26 human subjects, where 12 received active tcVNS and 14 received sham stimulation in tandem with acute traumatic stress. We collected continuous physiological signals related to the electrical and mechanical activity of the heart, peripheral blood volume, and respiration from electrocardiography (ECG), seismocardiography (SCG), photoplethysmography (PPG), respiratory effort (RSP) signals. Utilizing signal processing methods, we extracted parameters that are indicative of autonomic nervous system (ANS) activity, peripheral vasoconstriction, and respiration dynamics. Then, we employed machine learning techniques to determine the stimulation type, active or sham, from the extracted parameters and determined suitable sensing modalities for “in the wild” tcVNS use that could quantify therapy response, and ultimately enable closed-loop tcVNS delivery when a traumatic stress trigger is detected.

## II. Methods

### A. Human Subjects Study and tcVNS Application

The study focused on tcVNS effects on acute traumatic stress and was conducted under a protocol approved by the institutional review boards of the Georgia Institute of Technology, Emory University, and the Department of Navy Human Research Protection Office (HRPO). A total of 26 adults aged 18-65 who had experienced at least one psychologically traumatic event and based on Diagnostic and Statistical Manual-5 (DSM-5) criteria were recruited [34, 35]. Personal traumatic events were recorded upon recruitment and presented as voice recordings during the study session. After a baseline resting period, the stimulation was applied to the left side of the neck immediately after hearing the one-minute traumatic stress recording. The subjects kept supine position throughout the protocol. The protocol included two kinds of stimuli: active tcVNS or sham stimulus (GammaCore, ElectroCore, Basking Ridge, NJ), both of which were indistinguishable in appearance and operation with the following waveform characteristics: 5kHz sine bursts repeating at a rate of 25 Hz for the active tcVNS (0-30V peak output, adjustable intensity), and 0.2 Hz stepped pulse (0-14V peak output, adjustable intensity) for the sham stimulus, both lasting for two minutes. The subjects, the researchers, and the clinical staff were blinded to the device type. As the vagus nerve is typically located by the carotid artery, collar electrodes were placed on the left neck on top of the carotid pulse using a conductive electrode gel (GammaCore, ElectroCore, Basking Ridge, NJ) and the devices were operated by the researcher.

### B. Statistical Analysis

Subject demographics (age, gender, height, weight, body-mass index) and baseline physiological parameters were compared between the two device groups, active and sham, to understand whether the groups were significantly different from each other. Normality was assessed using Shapiro-Wilk test. The comparisons were made using t-tests for normal continuous variables, Wilcoxon rank-sum tests for non-normal continuous variables, and chi-squared tests for categorical variables. P-values lower than 0.05 were considered statistically significant. Table I summarizes the demographics and baseline physiological parameter information for the subjects, for each device group: active and sham. The groups were determined to be balanced with equivalent baseline characteristics, rendering the data amenable to use for further engineering purposes.

**TABLE I.** Demographics and Baseline Physiological Parameters

| Parameter | Active (n=12) | Sham (n=14) | P-value |
| --- | --- | --- | --- |
| Age [years] | 28.7 (6.8) | 30.8 (10.6) | 0.57 |
| Sex [F, %] | 5, 41.7% | 9, 64.2% | 0.25 |
| Weight [kg] | 76.8 (13.9) | 78.8 (14.1) | 0.86 |
| Height [cm] | 174.7 (11.1) | 170.5 (6.7) | 0.47 |
| BMI [kg/m<sup>2</sup>] | 25 (3.2) | 26.9 (4.2) | 0.37 |
| HR [bpm] | 67.4 (13.9) | 62.4 (9) | 0.63 |
| PEP [ms] | 66.5 (17.7) | 62.4 (9) | 0.98 |
| PPG Amplitude [a.u.] | 0.3 (0.3) | 0.1 (0.1) | 0.11 |
| PAT<sub>FOOT</sub> [ms] | 274 (31.7) | 261.2 (19.2) | 0.34 |
| PAT<sub>PEAK</sub> [ms] | 450.6 (38.2) | 437.1 (34.1) | 0.34 |
| RR [breath-pm] | 17.3 (5.4) | 18. (3) | 0.63 |
| RW [s] | 1.8 (0.7) | 1.6 (0.3) | 0.66 |
| RP [a.u.] | 0.8 (0.6) | 1.3 (0.9) | 0.27 |
| F = female; BMI = Body-mass index; bpm = beats per minute; breath-pm = breaths per minute, values indicate mean (SD). |  |  |  |

### C. Physiological Monitoring

ECG, SCG, PPG, RSP signals were collected continuously throughout the protocol using wireless amplifiers for 3-lead ECG, RSP, and transmissive PPG signals (Bionomadix RSPEC-R and PPGED-R, Biopac Systems, Goleta, CA). SCG was measured in the dorsoventral direction using a low noise accelerometer placed on the middle of the sternum (356A32, PCB Electronics, Depew, NY). RSP measurement was taken using a respiration belt and the PPG measurement was taken from the index finger. All data were simultaneously recorded using a 16-bit data acquisition system at 2kHz (MP150, Biopac Systems, Goleta, CA).

### D. Signal Processing and Parameter Extraction

Fig. 2a summarizes signal processing and parameter extraction paradigm, conducted in MATLAB (R2017b, Natick, MA). The physiological parameters related to ANS and peripheral physiological activity were extracted from four sensing modalities: ECG, PPG, SCG, RSP. First, the signals ECG, PPG, SCG were band-pass filtered with finite impulse response filters with cut-off frequencies 0.6-40Hz for ECG, 0.4-8Hz for PPG and 0.5-25Hz for SCG [36]. PPG and SCG signals were beat-by-beat segmented with beat lengths of 150ms and 600ms, respectively, with reference to the R-peaks of the ECG signal that are located by thresholding. These beat lengths were selected as they were adequate to capture the fiducial points in each beat. Exponential moving averaging was implemented for some of the parameter extraction tasks listed below to reduce motion artifacts [37]. The extracted parameters are listed below. *Heart rate (HR):* HR was extracted for multiple reasons. First, it is a basic vital sign used in clinical studies. Second, and more importantly, HR is regulated by both branches of the vagus-regulated ANS, the sympathetic (SNS) and parasympathetic nervous systems (PNS) [38]. Multiple implantable (direct) and noninvasive VNS studies noted changes in HR [23, 39, 40]. Therefore, instantaneous HR was computed from the R-R intervals of the ECG signals (in beats per minute) as a complex indicator of ANS balance.

**Fig. 2.**
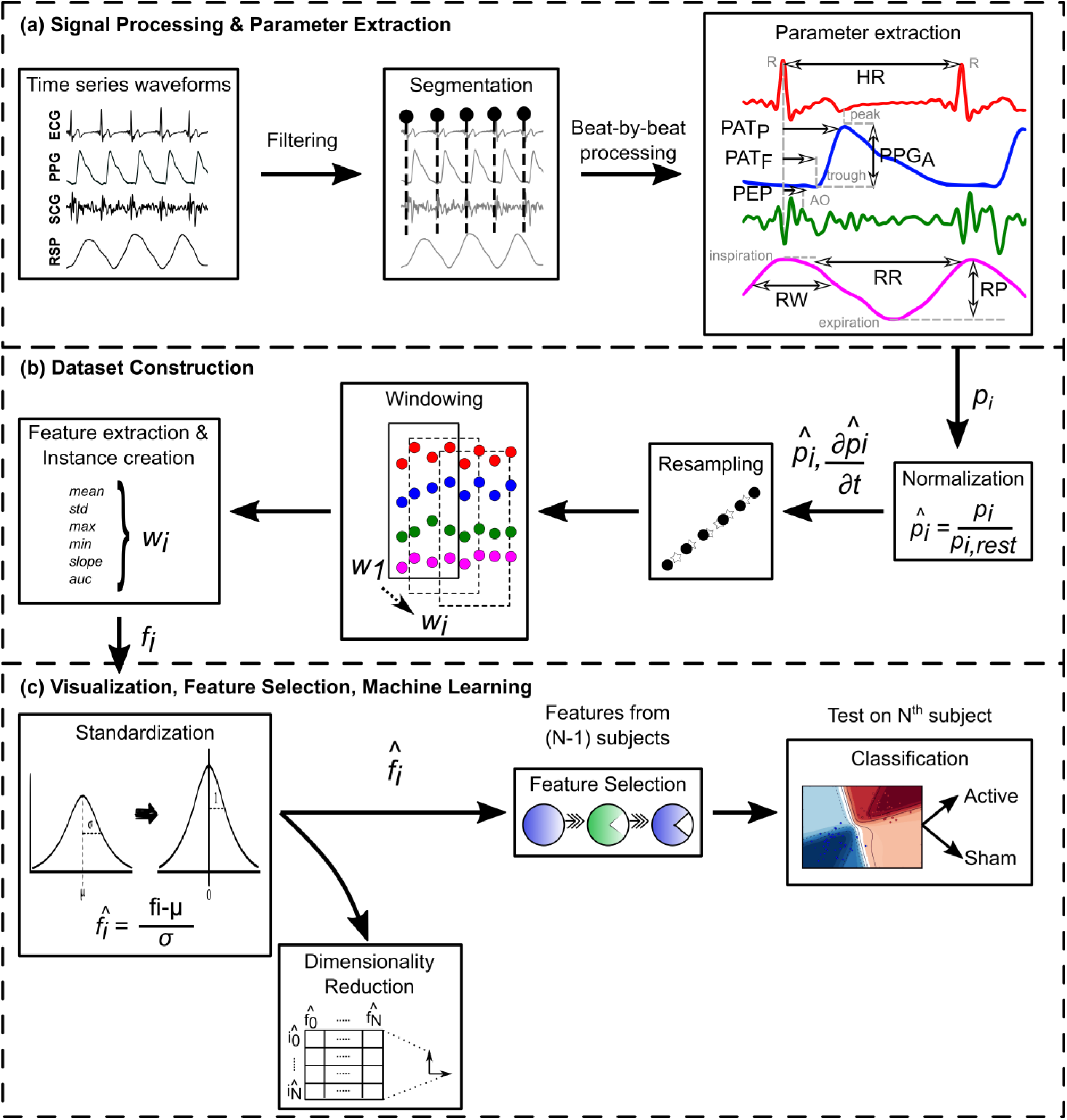
(a) ECG, PPG, SCG, RSP signals were processed and HR, PAT, PEP, PPG amplitude, RR, RW, RP were extracted as physiological parameters. (b) Using the extracted parameters, dataset constructed after normalization, resampling, and windowing. (c) After standardization, dimensionality reduction methods were applied for dataset visualization. Then, feature selection and machine learning were conducted. PAT_F_: PAT_FOOT_; PAT_P_: PAT_PEAK_; AO: Aortic opening; PPG_A_: PPG amplitude.

#### PPG amplitude and pulse arrival time (PAT)

The PPG signal is a rich and complex source of information about arterial tone (vasodilation or vasoconstriction), peripheral sympathetic activity, and relative changes in pulse pressure [41]; therefore, multiple parameters were extracted from this signal. The beat-by-beat amplitude of the PPG waveform, computed as the difference between the maxima and minima of each beat, was extracted as a measure of the amplitude of the peripheral blood volume pulse, and indirectly reflects sympathetic activity. PAT, a measure of relative changes in blood pressure and the pre-ejection period (PEP), was extracted as the time difference between R-peak of ECG to two different reference points of each PPG beat, named as PAT_FOOT_ (computed from through) and PAT_PEAK_ (computed from peak). A five-beat time constant for exponential moving averaging was used for PAT calculation.

#### PEP

PEP is a systolic time interval measured from ECG and SCG beats as an acute indicator of contractility and cardiac sympathetic activity [42]. It is computed as the time delay from ECG R-peak to the second peak in SCG signal, with a three-beat time constant for exponential moving averaging.

#### Respiratory measures

As the vagus nerve is heavily involved in parasympathetic control, we also extracted measures that indicate respiratory variations and are documented to dominate the parasympathetic function from the RSP waveform [43]. These measures include respiratory rate (RR), width (RW), prominence (RP). The RSP signal was detrended to avoid undesired DC offset using a sixth order polynomial for each one-minute signal segment. The rate of the inspiration peak appearance was extracted as RR (in breaths per minute). After finding the peaks, the width and prominence of each peak were also extracted as RW and RP.

### E. Dataset Construction

Fig. 2b depicts the dataset construction paradigm. The physiological parameters were extracted from stimulation and post-stimulation intervals, which correspond to the data from the last minute of stimulation and one-minute data from three minutes after the stimulation ended, respectively. Then, these parameters were divided by the mean values obtained from the rest period to normalize inter-subject variability, as in our prior physiological sensing work [44]. The rest data were obtained before the stimulation protocol started, at the same body position the stimulation was carried out. The dataset for machine learning was constructed from the time-series data obtained from the normalized intervals. The extracted parameters were resampled to the length of the parameter that has the maximum length to equalize the length of each parameter within an interval for further usage of the resampled data in windowing (to obtain equal length of arrays for windowing), using an antialiasing FIR lowpass filter and compensating for the delay introduced by the filter [45]. Resampling was necessary as the length of respiratory features are approximately half of the beat-by-beat features per unit time, as human respiration and heartbeat frequency are different (12-20 breaths per minute respiratory rate versus 70-100 beats per minute heart rate). Then, instances were created by using 10-sample sliding windows with 9-sample overlap (90% overlap). The features consisted of the mean, standard deviation (std), maximum (max), minimum (min), area under curve (auc) and slope (slp) of the extracted physiological parameters in each window. The approximate first order derivatives (differences between the adjacent elements) were also computed to generate additional parameters. The same feature calculation methodology was applied to the difference matrices (except slope). The multi-dimensional feature matrix was constructed from all extracted features and the corresponding labels as device types (tcVNS, sham). This matrix included features as columns and instances as rows, consisting of 176 features and 1780 instances (827 tcVNS, 953 sham). Overall, these features were derived from 16 time-series data (HR, PEP, PPG Amplitude, PAT_FOOT_, PAT_PEAK_, RR, RW, RP in two intervals), with 11 mathematical properties derived from each of them and their differences. Later, the data were standardized across each feature to have a mean of zero (subtracting the mean in the numerator in Fig. 2c) and a standard deviation of one (unit variance, by dividing by the standard deviation of the data as seen in Fig. 2c). Standardization is necessary because some parameters respond differently for the same level of stress due to the physiological nature of the measurement. (i.e., 5% decrease in PEP or 80% decrease in PPG amplitude might both indicate similar levels of stress).

### F. Visualization, Feature Selection, Machine Learning

Fig. 2c summarizes the visualization, feature selection, and machine learning approach. For demonstration purposes, t-distributed Stochastic Neighbor Embedding (t-SNE) was applied to the high-dimensional standardized feature matrix to visualize the dataset in two dimensions [46, 47]. For the machine learning paradigm, our primary concerns were 1) minimizing the number of features/sensing modalities used in classification to optimize the computational power and to determine which sensing modalities are necessary for a wearable implementation (note that there are a total of 176 features from four sensors with unknown contributions to the classification task), 2) developing a realistic validation paradigm for a new subject who has just started undergoing stimulation. To address 1, we implemented a univariate feature selection paradigm based on sorting ANOVA F-statistics of features to eliminate redundant data without incurring significant loss of information [48]. Note that this feature selection step is not mandatory but optional. Assuming no need for optimization, not applying this step (simply using all features) did not cause a dramatic decrease in performance (See Table II, SVM with sigmoid kernel, which was used as our classifier). To address 2, we wanted to ensure that the training dataset are not biased for a new, incoming subject by using leave-one-subject-out cross validation (LOSO-CV). Typical methods such as K-fold cross validation would not guarantee that the training dataset does not have data from the incoming subject. The fact that there is data from the incoming subject in both the training and testing datasets will likely make the model know more about the target subject than it should. With an entirely new subject (not in the training dataset), the K-fold trained model will potentially perform poorly because it did not include data from the new subject in the training set before. LOSO-CV, on the other hand, guarantees there is no data from the incoming subject in the training dataset in model evaluation process already, hence this is a conservative approach to assess an expected performance for an incoming subject. Feature selection and LOSO-CV were implemented as follows: For each LOSO-CV loop, one subject was left out of the feature selection and classification model training, then used for testing. The procedure was repeated for each of the 26 subjects. We determined the number of top features by implementing the feature selection and LOSO-CV and swiping the number of top features from 1 to 176. For each case, we computed the area under curve obtained from the receiver operator characteristics (ROC AUC) and plotted k_topfeatures_ versus ROC AUC. Then, we determined a range for k_topfeatures_ where ROC AUC is high (>0.9) and robust enough to ensure good performance and k_topfeatures_ is low enough to reduce processing time.

**TABLE II.** ROC AUC Scores of Classifiers Derived Using LOSO-CV Using All Features

| Naïve Bayes | Random Forest | k-NN | Logistic Regression | MLP |
| --- | --- | --- | --- | --- |
| 0.52 | 0.76 | 0.56 | 0.80 | 0.71 |
| Decision Trees | SVM (Linear) | SVM (Polynomial) | SVM (RBF) | SVM (Sigmoid) |
| 0.62 | 0.80 | 0.66 | 0.65 | 0.98 |

ROC AUC = Receiver operator characteristics area under curve; k-NN = k-Nearest Neighbors; MLP = Multilayer Perceptron; SVM (Kernel) = Support Vector Machine (Kernel Function); RBF = Radial Basis Function.

Following the determination of feature selection and the needs for model evaluation, we explored the most effective classification architecture for predicting stimulation presence from physiological signals. Using LOSO-CV, we used the following classifiers: Naïve Bayes (for predicting baseline performance), Random Forest, k-Nearest Neighbors (k-NN), Logistic Regression, Multilayer Perceptron, Decision Trees, and Support Vector Machine (SVM) with linear, polynomial, radial bases function (RBF), and sigmoid kernels. Table II (see Results section below) lists the ROC AUC values for each classifier. We observe that SVM with a sigmoid kernel performs the best among different classifiers used, hence we decided to use this classifier.

## III. Results

We performed multiple classification runs with a sigmoid SVM classifier using features from different sensors alone and their combinations. Figure S1 shows the number of features versus ROC AUC, plotted from one feature to the maximum number of features based on the sensors chosen. It is apparent that ROC AUC>0.8 could be obtained from different sensor combinations.

By using the described feature selection and machine learning methodologies, we obtained the highest ROC AUC (>0.90) and highest accuracy, precision, recall (>0.90) for four cases: By using: 1) all sensors, at least 140 top features out of 176 (Figure S1-i); 2) ECG and PPG sensors, at least 70 top features out of 88 (Figure S1-vi); 3) ECG, SCG, and PPG sensors, at least 99 top features out of 110 (Figure S1-x), and 4) ECG, PPG, and RSP sensors, at least 84 features out of 154 (Figure S1-xi). As our aim was to minimize the number of sensing modalities and features to reduce complexity for a future wearable implementation, we focused on the classification results for the second case, the combination of ECG and PPG. The features in this dataset contain HR, PPG amplitude, and PAT.

Our next goal was to analyze the results obtained from ECG and PPG. Fig. 3 details the outcomes for the features obtained from ECG and PPG. Fig. 3a shows the dimensionality reduction (t-SNE) plot grouped by tcVNS and sham clusters. A nonlinear separation exists between the device groups. Fig. 3b shows how the number of selected top features change the ROC area. A ROC area of >0.9 can be obtained by using 70 top features out of 88 (and beyond). Fig. 3c-d summarize the machine learning outcomes for the classification using top 71 features out of 88 (note that outcomes are similar between 70 to 88). The classification resulted in 25 correctly classified subjects out of 26 total subjects (12 active tcVNS, 14 sham) with LOSO-CV. There was one false negative subject. Macro-averaged accuracy, precision, recall, and F-1 scores of 96% with 0.96 ROC AUC were obtained as the performance outcomes. Due to the feature selection method applied in each LOSO-CV loop, the features used in the classification slightly differed for each subject. Fig. 4 shows the top 5 features obtained by applying the feature selection method to the whole dataset of 88, sorted by ANOVA F-values. PPG and HR features resulted in the largest F-values. Lastly, we calculated the time complexity (the time elapsed encompassing all tasks ranging from signal processing to machine learning) for each subject as measured on a Core i7-6500U CPU @2.5GHz 12GB RAM personal laptop. We found that 2.6 (1.1) seconds (mean (SD)) were required to generate an output class for an incoming subject.

**Fig. 3.**
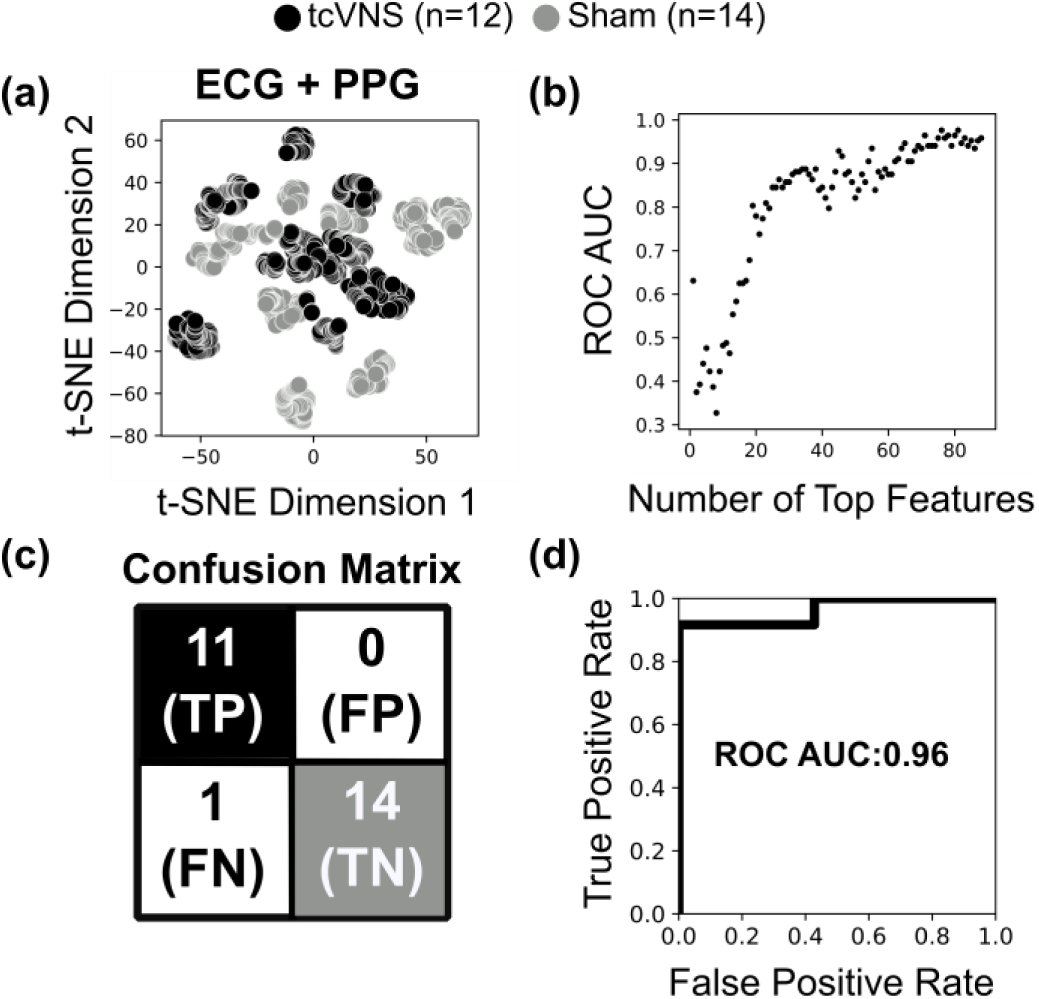
Dimensionality reduction and classification outcomes for separating the stimulus types: active tcVNS and sham. (a) Dimensionality reduction applied to the high-dimensional feature matrix using t-SNE constructed from features from ECG and PPG. (b) Number of Top features selected using ANOVA F-score-based feature selection versus receiver operator characteristics (ROC) area under curve (AUC). ROC AUC is robust to Top Features from 70 to 88. c) Confusion matrix for the classifier, obtained with LOSO-CV and minimum number of features (71). (d) Receiver operator characteristics (ROC) for the classifier. A ROC area under curve (AUC) of 0.96 was obtained. Classification outcomes vary minorly with Top Features from 70 to 88.

**Fig. 4.**
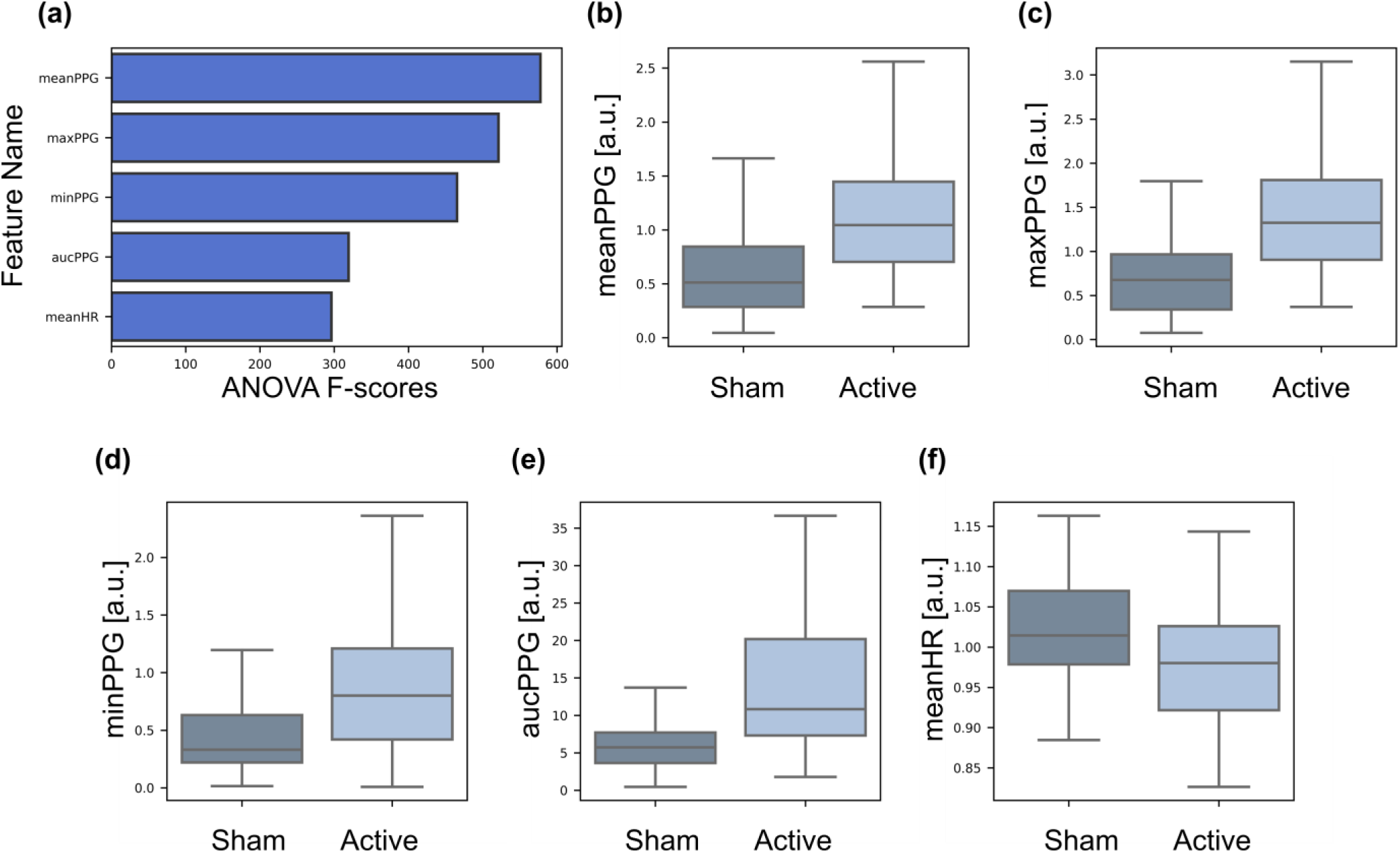
Top 5 features sorted by ANOVA F-values and their boxplots grouped by sham and active tcVNS classes. The top features were calculated from the full feature set obtained from ECG and PPG sensors.

## IV. Discussion

In this work, we investigated methods for detecting target engagement for tcVNS by using cardiovascular and peripheral signals from a double-blinded study. The dataset constructed from ECG and PPG signals yielded sufficient information to detect whether therapy response occurred (active device), resulting in 25 correctly classified subjects out of 26. The false negative subject (active subject classified as sham) was a female aged 24 years. The combination of other sensing modalities resulted in similar classification outcomes, slightly changing the confusion matrix. Single sensors/features do not offer the same performance as the combinations. Thus, there is not a single biomarker or pattern that could be easily recognized by the human eyes; there is likely a complex, high-dimensional relation between multiple features that requires multiple modalities.

The investigation of the most salient features reveals that features related to PPG and HR perform favorably compared to others. HR is regulated by both SNS and PNS activity [38]. Stimulation of the vagus nerve typically decreases SNS activity or increases PNS activity, also observed in the clinical portion of this study [30]. Moreover, increase in PPG amplitude was noted when tcVNS was paired with traumatic stress, compared to sham. From Fig. 4b-f, PPG features increased and mean HR decreased for the tcVNS group. Both of these changes indicate a decrease in sympathetic tone, which perhaps makes the separation between the classes possible by these features.

The analysis of machine learning performance outcomes is essential to gain an understanding regarding the translation of the methods to at-home settings: with our classifier, the accuracy, precision, recall, F-1 scores were 0.96. Thus, the classifier could provide a binary (e.g., red or green) indicator to the user following stimulation regarding whether the nerve target was successfully engaged. Among 100 stimulation administrations upon traumatic stress triggers, we would be able detect whether the stimulation happened correctly 96 times.

The methods described in this study leverage physiological signals that are convenient to obtain with wearable sensing modalities, such as the smartwatches that measure ECG and PPG. The framework presented herein could remain the same and could be generalized to include other types of sensory data. Note that although the measured computation time was 2.6 seconds on average to generate an output class for an incoming subject, this was computed with a personal laptop. A final implementation of this pipeline would not require a fully embedded platform as no precision timing is necessary. The collected signals could be transferred to a Cloud-based distributed system that employs parallel processing capabilities to further reduce this 2.6-second computation time if needed.

Among the modalities we used, HR and respiratory effort information are typically collected in clinical practice, in a non-continuous manner. Therefore, we explored using only these features. Using the features obtained from HR (Figure S1-ii), RSP (Figure S1-v), and their combination (Figure S1-viii) for the classification task do not outperform the reported results.

Other measures that might indicate stimulation presence from the literature are the use of microneurography, pupil size, serum cytokines (anti-inflammatory effects as quantified by tumor necrosis alpha, interleukin-6), heart rate variability (HRV), or evoked potentials resulting from stimulation [10, 20, 49]. Microneurography and serum cytokines are not non-invasive, not pursuant to the goal of real-time identification, and are confined to clinical settings, electroencephalogram [50] requires bulky multi-channel equipment, while pupil size was not found to give favorable results for stimulation in prior work. As for HRV, this measure does not have a continuous nature and requires a long-term clean recording to compute a single number based on the variability in R-R intervals. An auricular stimulation study noted changes in HRV with long-term recordings [10], while some studies did not note changes in HRV [17, 51].

A number of limitations exist for the study described here. First, this study employs a reactive approach. Traumatic stress requires a flashback occurrence: the subjects should remember their memories to be stressed, which requires some time starting from the introduction of the traumatic recording. We applied stimulation immediately after the traumatic stress recording as our prior brain imaging studies demonstrated that the arousal persists following rumination of the traumatic script recording [52, 53]. Future studies should consider sweeping the timing of the stimulation, which might possibly downregulate the autonomic reactivity even before reaching a peak stressed state, based on prior preclinical studies [54, 55]. Second, this study did not have continuous blood pressure recordings, hence we could not use blood pressure-related measures except pulse arrival time (PAT) obtained from ECG and PPG, which is a measure related to *both* continuous blood pressure and cardiac contractility [56]. Nevertheless, analyses on the effects of auricular or cervical stimulation on cardiovascular and autonomic function have produced mixed outcomes throughout many studies that use basic vital signals such as HR, HRV, or BP [10, 17, 18, 21, 23, 51], hence these basic measures are not likely to be sufficient for monitoring stimulation presence. Lastly, in the current study, we obtained a transmissive PPG signal that requires the photodiode (PD) and light emitting diode (LED) combination to be at the opposite sides of the skin. A reflective PPG sensor (both PD and LED on the same side of the skin) might be more appropriate for a comfortable and minimally obtrusive wearable device.

## V. Conclusion

This study demonstrates the first effort to provide real-time inputs that could be used in tcVNS therapy response, and further for closed-loop modulation for at-home tcVNS technologies. Multimodal signal fusion might be a viable approach in determining whether the stimulation occurred as expected. In addition to the investigation of individual parameters, sensor fusion could be instrumental in translating tcVNS to unsupervised settings to improve therapy response in a home-based setting. ECG and PPG sensing appear to provide relevant information regarding the stimulation delivery, and both signals could be obtained noninvasively, and are also prevalent in commercially available wearable sensing devices. The methods presented herein could thus be deployed in wearables allowing for a convenient home-based approach to supporting accurate and effective delivery of noninvasive vagal nerve stimulation therapies.

Future studies should focus on the following to facilitate successful translation to clinical practice. Continuous blood pressure should be measured as an additional physiological parameter to be studied as it includes important vascular information; effects of the time of stimulation (before or during traumatic stimuli) on efficacy should be determined; and the ability of indices derived from reflective PPG sensors to capture the key physiological information should be quantified. Additionally, stimulation induces parameter-specific changes in physiology [11]. Therefore, regression models of stimulation parameters onto the changes in cardiovascular and peripheral measures could provide a better understanding for inter-patient variability in noninvasive VNS studies.

## Data Availability

The data and MATLAB/Python codes, that support the findings of this study, are available from the corresponding author (Nil Gurel) upon reasonable request.

## Acknowledgments

This work was supported by the Defense Advanced Research Projects Agency (DARPA), Arlington, VA, under Cooperative Agreement N66001-16-2-4054. Dr. Shah was sponsored by the National Institutes of Health, Award K23 HL127251.

